# Multi-omic triangulation identifies molecular candidates of atopic dermatitis severity

**DOI:** 10.1101/2025.08.04.25332125

**Authors:** Katie Watts, Matthias Hübenthal, Silke Szymczak, Hannah Cherry, Heming Xing, Antonio Federico, Oxana Lopata, Marcio Acencio, Manuela Hofner, Samuel Lessard, Thomas Battram, Maris Teder-Laving, Laurent Thomas, Karin Ytterberg, Aditya Krishna, Ben Brumpton, Jan Hartmann, Ravi Ramessur, Jake Saklatvala, Sarah Watkins, Wouter Ouwerkerk, Maritza Middelkamp Hup, Clement Chatelain, Emanuele De Rinaldis, Johannes Kettunen, Laura Huilaja, Shameer Khader, Zhipeng Liu, Bailin Zhang, Katherine Klinger, Kaisa Tasanen, Kristian Hveem, FinnGen, Estonian Biobank research team, Chengliang Dai, Alena Buyx, Marie-Christine Fritzsche, Sinead Langan, Nora Langreder, Stefanie Eyerich, Ellen Van den Bogaard, Ilka Hoof, Paola Lovato, Karen Page, Erin Macdonald-Dunlop, Angela L-A Bosma, Lukas Roth, Sandro Bruno, Frank Kolbinger, Külli Kingo, Jochen Schmitt, Thomas Werfel, Bo Jacobsson, Pol Solé-Navais, Mari Løset, Alexandra Hicks, Venkata Satagopam, Josine Min, Nick Dand, Stephan Weidinger, Dario Greco, Klemens Vierlinger, Marek Ostaszewski, Nanna Fyhrquist, Rowann Bowcutt, Soumyabrata Ghosh, Joe Rastrick, Sara J Brown, Catherine Smith, Lavinia Paternoster

**Author notes:** joint senior authors (Sara J Brown, Catherine Smith, Lavinia Paternoster).

## Abstract

Atopic dermatitis (AD) is a common skin disease with most of the health, social and economic impact driven by those with more severe disease. Determining the molecular pathways that influence severity is therefore crucial, offering opportunity to identify novel drug targets, as well as use in risk prediction tools. In this large-scale multi-omics study, we used complementary methods and datasets to identify molecular markers with robust evidence for involvement in AD severity. We undertook a case-only genome-wide association study meta-analysis (N=100,766) and subsequent transcriptome-wide association study (TWAS), differential expression meta-analysis in blood (N=340) and skin (N=185) as well as a differential protein abundance analysis in blood (N=75). A total of 440 genes/proteins showed evidence of association across all the analyses. Of these, four were significant in two or more analyses. For *CEP85* (P_expression_=2.8×10-7; P_TWAS_=8.2×10-13), a gene not previously associated with AD, we also found strong evidence that the genetic variants affect *CEP85* mRNA expression in monocytes. Functional in vitro follow-up showed that *CEP85* over-expression in monocyte-derived macrophages can disrupt phagocytosis which we hypothesise may contribute to severity by impairing phagocytosis of *S aureus*. Together this work provides evidence of genetic risk and candidate molecular pathways to severe AD.

## Introduction

Atopic dermatitis (AD) is an inflammatory skin disorder that affects up to 10% of adults and an estimated 130 million globally^1^. It is clinically heterogeneous, with variability in the severity and trajectory of disease^2^. Approximately 40% of people with AD can be classified as having moderate or severe disease^3–5^ and disease severity has been shown to correlate with worsening quality of life, multimorbidity and healthcare costs^6^.

While there have been major advances in the understanding of disease susceptibility and the underlying disease biology, there has been less attention on the mechanisms underpinning more severe disease. Identifying genetic and molecular markers associated with severe disease is important as it potentially provides novel drug target opportunities for the highest need population. It may also enable the development of molecular tests to predict and/or monitor disease trajectories for patients which will enable a more proactive approach to disease management. Although studies have investigated various biomarkers for association with AD severity ^9–25^, a systematic, multi-omic approach has not yet been performed. There have only been a limited number of studies investigating the genetics of AD severity^26–28^, with the only established genetic makers being *FLG* null variants. Without supporting genetic evidence, it is also uncertain whether reported biomarkers from other omics have any causal impact.

One challenge which has limited the wide-scale study of severity is in the definition and capture of cases. Disease severity may be recorded at a point in time or over a period of time and relates not only to inflammation in the skin itself but also to the impact (such as symptoms, quality of life and health care use)^7^. There are well established tools to capture the extent and degree of inflammation in the skin, such as the Eczema Area and Severity Index (EASI) and SCORing of Atopic Dermatitis (SCORAD). However, such validated eczema severity scores are not routinely collected in large-scale population cohorts. Even in the smaller patient-focused cohorts where severity measures are available, mapping between different methods is not straightforward. However, as we have recently demonstrated in the BIOMAP consortium, disease severity can be studied in population cohorts through use of clinically recorded proxy measures such as healthcare use^7^. It is also possible to map between EASI and SCORAD/o-SCORAD to achieve harmonised severity scores, making meta-analysis of studies that use different measures now viable^8^.

In the BIOMAP consortium we set out to identify causal mechanisms for disease severity in large-scale meta-analysis, across multiple ‘omics layers’ (genome-wide association analysis of genetic variants, gene expression levels in blood and skin, and proteomics in serum). We used a triangulation approach, whereby a set of complementary methods and datasets are brought together to address the same underlying causal question^29^, to identify molecular markers with the most evidence for involvement in AD severity.

## Results

We have conducted a series of analyses including a genome-wide association analysis (GWAS) of genetic variants, differential gene expression (DGE) analyses (in blood and skin) and a proteomic abundance analysis in serum. An overview of the multi-omic analyses and results is given in **Figure 1**. The complete summary statistics for all the analyses alongside data visualisation tools are available via our dataviz platform (see data availability).

**Figure 1.**
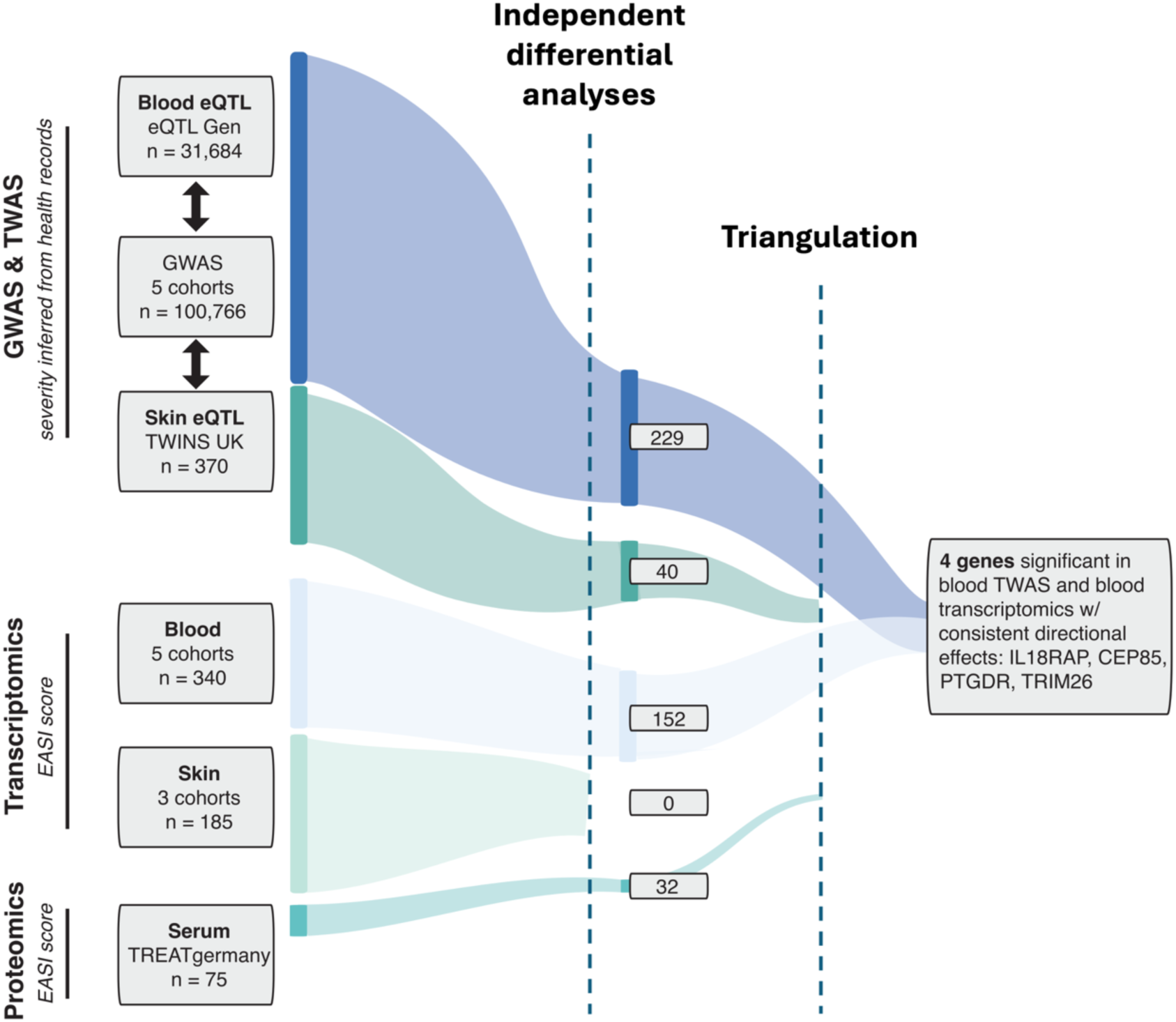
Flow diagram showing multi-omic triangulation method. We performed large scale meta-analyses to identify molecular biomarkers associated with atopic disease severity. These were performed for multiple omics; genome-wide association analysis (GWAS) of genetic variants, gene expression differences (in blood and skin) and proteomics differences in serum (**Supp Table 1**). To convert the GWAS results into gene-level results suitable for triangulation, we performed two transcriptomic-wide association studies (for blood and skin) using publicly available eQTL datasets. Significant TWAS genes were those that passed a Bonferroni corrected p-value threshold of 3.65×10^-6^. Pearson’s correlation was then used to identify the number of independent genes (R^2^ > 0.5). For transcriptomics, differentially expressed genes were those with evidence (p_FDR_<0.05) of correlation with EASI score across studies described in **Supplementary Table 1**. For proteomics, differentially abundant proteins were those with evidence (p_FDR_<0.05) of correlation with EASI score in the TreatGermany cohort. Triangulation was performed by identifying genes that were significant at thresholds corrected for multiple testing in >1 omics. In the case of overlap between TWAS and gene expression results, we selected only genes with consistent directions of effect.

### Genome-wide association meta-analysis identifies 13 AD susceptibility loci to be also associated with AD severity

The GWAS meta-analysis included 100,766 individuals with AD, classified as having severe disease (or not) based on clinical records of hospitalisation and/or use of certain potent treatments (as described in <u>methods</u>). In total, 25,744 individuals were categorised as having severe AD and 75,022 were classified as non-severe AD for comparison (**Supplementary Table 1**). There was no evidence of genomic inflation in either the individual studies (λgc < 1.02) or the overall meta-analysis (λgc = 1.01).

The meta-analysis found two loci that were genome-wide significant (P < 5.0×10^-8^, **Figure 2**), both of which are known signals for susceptibility. The strongest signal was for *FLG* null mutation 2282del4 (P = 2.5×10^-12^, OR = 1.44 95% CI =1.3-1.6), a well-characterised variant that among other *FLG* null variants are the only established markers of AD severity^30^.

**Figure 2.**
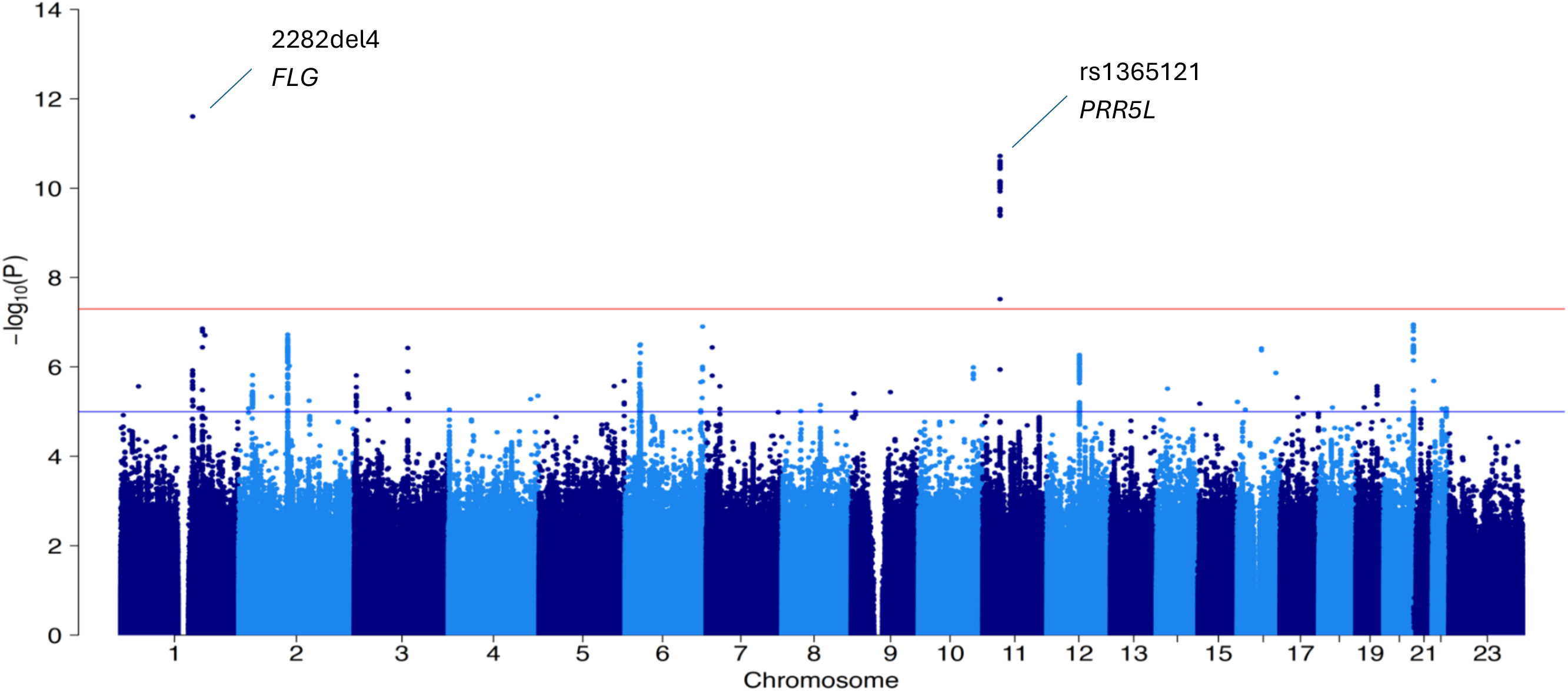
Manhattan plot of atopic dermatitis severity. Meta-analysis of 5 cohorts, N=100,766 AD patients. Annotated are the 2 significant loci from GWAS.

The second significant locus resides on chromosome 11p13 with the lead variant being rs1365121 (P = 3.5×10^-12^, OR = 1.2, 95% CI = 1.13-1.25, **Supplementary Figure 1**). Variant to gene mapping analyses prioritised *PRR5L, TRAF6, COMMD9* and *LDLRAD3* as possible causal genes (**Supplementary Table 2**). In particular, there was strong evidence from expression quantitative trait loci (eQTL) in blood for *PRR5L* (posterior probability for genetic colocalisation between AD severity and *PRR5L* expression signals (PP.H4) = 99%). Fine-mapping analyses narrowed the 95% credible set to 20 variants, none of which are protein coding (**Supplementary Table 3**), but all are intronic to *PRR5L*. As both genome-wide loci have previously been reported for susceptibility, we then explored other known susceptibility loci. Of the 81 lead variants associated with AD susceptibility from Budu-Aggrey (2023)^31^, 13 were also associated with severity after multiple testing correction (p<6.2×10^-4^, **Supplementary Table 4**).

### Transcriptome-wide association study finds gene expression levels in blood and skin associated with AD severity

To enable triangulation with the other molecular layers of evidence, we performed transcriptome-wide association studies (TWAS), a post-GWAS gene prioritisation approach that leverages eQTL data to detect significant gene-trait associations. Here, we used our full GWAS summary statistics and eQTL data from blood (eQTL Gen) and skin (TWINS UK) as input.

This identified 513 genes in blood and 40 genes in skin that were significantly associated at thresholds corrected for multiple testing (**Supplementary Table 5**). We tested for non-independence between significant genes and found 229 independent blood TWAS signals while all 40 skin gene signals were already independent.

### Differential gene expression analysis identified 152 genes in blood associated with AD severity (EASI)

We next performed two DGE meta-analyses (one in blood and one in skin) using a series of independent cohorts with severity scores and transcriptomics data. For all cohorts we adjusted for age, sex and surrogate variables.

In blood, DGE meta-analysis of three cohorts (N=340) identified 152 genes associated with our reference standard measure of overall skin disease severity (predicted EASI - see methods) p_FDR_<0.05 (**Supplementary Table 6**). Of these, 98 genes were upregulated and 54 were downregulated in patients with higher predicted EASI scores (**Figure 3**).

**Figure 3.**
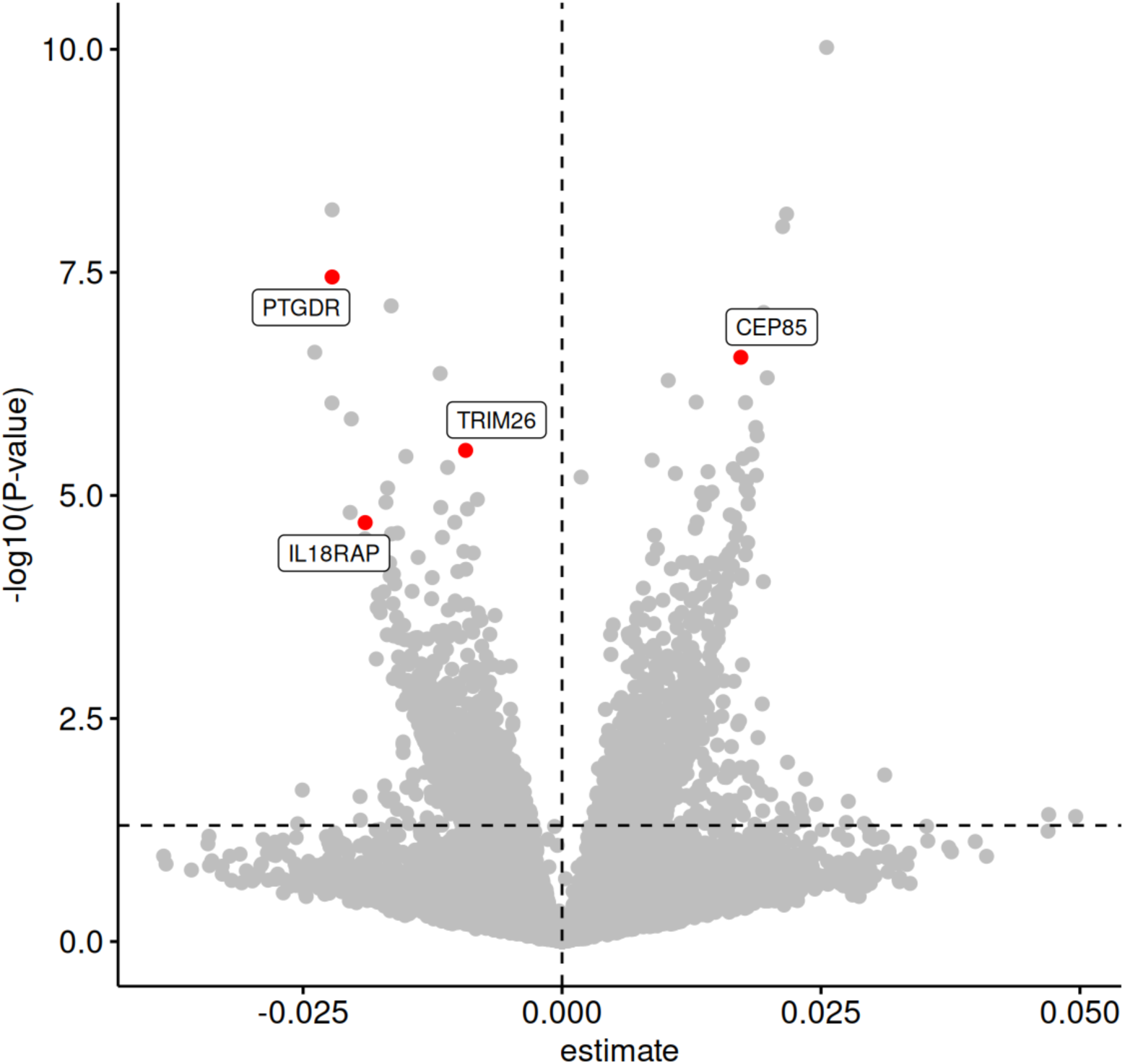
Genes significantly associated with predicted EASI score in blood. The horizontal dashed line shows a p-value threshold of 0.05. The 4 genes with significant evidence in a second omic analysis are labelled.

In skin, DGE meta-analysis of five cohorts (N=185) did not identify any genes that met our threshold for association with predicted EASI, p_FDR_<0.05.

In sensitivity analyses, we identified potential interactions between predicted EASI and sex/age for 4 genes that we later discuss in detail (**Supplementary Figure 2**), whereby the strength of the relationship between gene expression and EASI appears to increase with age. However, this analysis was underpowered which is demonstrated by the overlapping confidence intervals across the strata.

### Differential serum protein abundance analysis identified 32 proteins associated with AD severity (EASI)

For one of the cohorts contributing to the expression analysis (TreatGermany N=75), serum protein levels were also available for 2,174 proteins from high-throughput mass-spectrometry proteomic profiling. Using these data, we identified 32 proteins whose abundance associated with EASI score (FDR corrected *p* < 0.05; **Figure 4, Supplementary Table 7**). Of these 28 were upregulated and 4 were downregulated in individuals with increasing disease severity (EASI). QIAGEN Ingenuity Pathway Analysis (IPA) revealed that of the significant proteins, IL-4 is a significant upstream regulator of 9 (GSR, KCTD12, LDHA, LTA4H, PKM, POSTN, TNC, DSC1, GLIPR2, **Supplementary Figure 3).**

**Figure 4.**
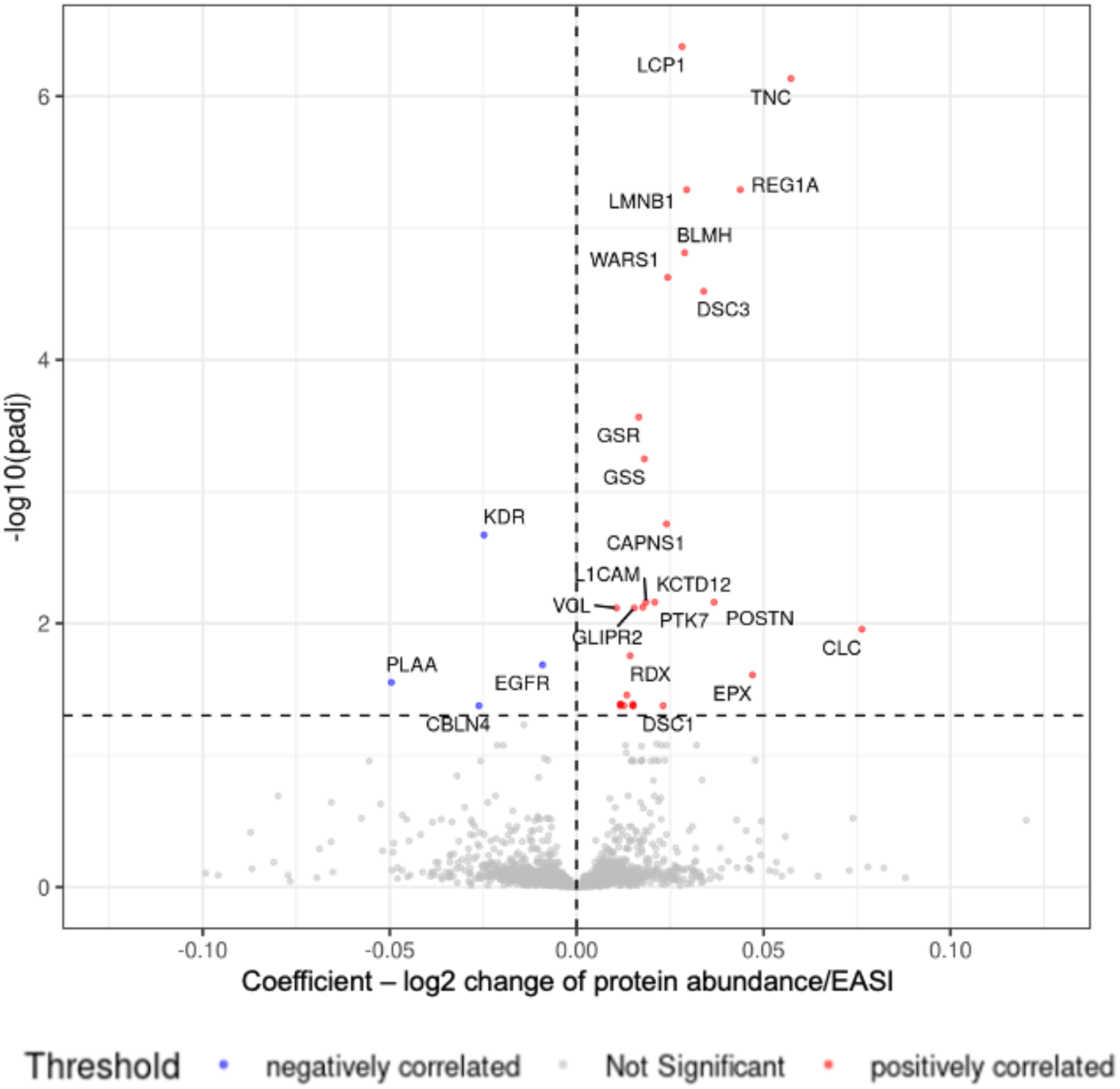
Proteins significantly associated with EASI score (TREATGermany N=75) in serum. Protein abundance was measured by mass-spectrometry. Proteins significant at an FDR corrected p-value threshold of 0.05 are labelled.

### Combining evidence across omics analyses implicated relevant biological mechanisms

In total 440 independent genes had evidence of association with AD severity in at least one of the omics analyses. To assess if these genes have known biology for AD susceptibility, we interrogated the AD disease map^32^ (a manually curated resource generated through literature review of pathways and mechanisms related to molecular mechanisms of AD). We found that only 14 of the 440 were present on the AD map which we highlight on a ‘eczema severity biomarkers’ map overlay, viewable here: https://bit.ly/admap_for_reviewers. We also investigated if any of our genes had literature-supported connections to genes present on the map. This identified a further 77 genes with connections to the AD map (29 with evidence from 10 or more sources) (**Supplementary Table 8**) but reduced to 30 when filtering the connections for DisGeNET^33^ conditions annotated with “skin diseases” or “inflammatory diseases” (**Supplementary Table 9**).

The results from all three blood-based omics analyses (and their interacting proteins) were then mapped onto a co-expression network to generate an extrapolated community of 2426 genes that we here call a “severity module” (See methods). Pathway enrichment analysis of the module found 482 enriched pathways (FDR<0.05, **Supplementary Table 10**). The most enriched pathways were related to generic biological mechanisms, but other significant pathways (after FDR correction) were related to antigen presentation and interleukin signalling. Despite the fact we found little evidence for individual skin biomarkers, the skin-based pathway enrichment analysis (with a “severity module” of 1656 genes) identified 374 pathways that were enriched (FDR < 0.05). Similarly to blood, the most enriched pathways were generic biological pathways, but other significant pathways did include those related to T-cell signalling and activation (**Supplementary Table 11**), which may be more relevant to AD.

### Assessment of biomarkers previously reported to be associated with severity

Several genes/proteins have previously been reported to be associated with AD severity. We performed lookups in our data of those that had been previously reported in 2 or more studies (**Supplementary Table 12**). Of the 12 included, only BLMH replicated in the same omic (proteomics) and tissue (serum) as the discovery. However, it is important to note that many of the genes/proteins were not included in our most relevant analysis/tissue (due to technology differences in profiling).

### Four genes showed evidence across multiple omics layers in triangulation analysis

Notably, we found four genes (*CEP85, IL18RAP, PTGDR and TRIM26*), with evidence (with consistent direction of effect) from more than one omics layer (**Figure 1**, **Table 1**). These four were not detectable in the proteomics mass-spectrometry profiling and so all were supported by both gene expression and genetic evidence. *IL18RAP* appears in our previously generated AD disease map and both *PTGDR* and *TRIM26* interact with genes in the map (1 and 4 links, respectively, **Supplementary Table 13**).

**Table 1.** Four molecular candidates with consistent evidence across multiple omics.

| Gene | Blood |  |  |  |  |  | Skin |  |  |  |  |
| --- | --- | --- | --- | --- | --- | --- | --- | --- | --- | --- | --- |
|  | Transcriptomics |  |  | TWAS |  | Proteomics | Transcriptomics |  |  | TWAS |  |
|  | Beta (SE) | p-value | FDR P-value | Best Z-score | p-value | p-value | Beta (SE) | p-value | FDR p-value | Best Z-score | p-value |
| *CEP85* | 0.017 (0.003) | 2.80x10<sup>-7</sup> | 8.10x10<sup>-4</sup> | 12.5 | 8.20x10<sup>-13</sup> | *Not measured* | 0.002 (0.005) | 0.65 | 0.99 | -3.0 | 0.19 |
| *PTGDR* | -0.022 (0.004) | 3.60x10<sup>-8</sup> | 1.84x10<sup>-4</sup> | -9.1 | 6.20x10<sup>-7</sup> | *Not measured* | -0.03 (0.006) | 1.90x10<sup>-3</sup> | 0.99 | 2.6 | 0.43 |
| *IL18RAP* | -0.019(0.005) | 2.00x10<sup>-5</sup> | 1.04x10<sup>-2</sup> | -11.8 | 2.60x10<sup>-11</sup> | *Not measured* | 0.002 (0.005) | 0.65 | 0.99 | -3.3 | 1.5x10<sup>-3</sup> |
| *TRIM26* | -0.0093 (0.002) | 3.10x10<sup>-6</sup> | 4.26x10<sup>-3</sup> | -19.9 | 9.20x10<sup>-31</sup> | *Not measured* | -0.012 (0.005) | 2.60x10<sup>-2</sup> | 0.99 | -6.6 | 0.07 |

The results for the 4 genes with significant evidence of association in 2 or more omics are shown. For the transcriptome wide association study (TWAS), a Bonferroni corrected p-value threshold of 3.65x10<sup>-6</sup> was used to determine significance. For transcriptomics (differential gene expression analysis), FDR was used to correct the p-values. All 4 genes were not detectable in the proteomics mass-spectrometry profiling.
SE = Standard error of beta coefficient .

#### CEP85

*CEP85* gene expression in blood was positively associated with AD severity (beta = 0.017, SE = 0.003, p_FDR_ =8.1×10^-4^) and the independent blood TWAS suggested a causal role of this gene (Z-score = 9.3, p = 8.3×10^-13^) (**Figure 5**). This was supported by Mendelian Randomisation (MR), which showed a causal effect of *CEP85* expression in blood on AD severity (beta = 0.08, SE = 0.027, p_IVW_ = 2.4×10^-3^). Investigating immune and skin cell types, we found the GWAS signal colocalises with *CEP85* eQTL signal in monocytes in three different eQTL datasets^34–36^ (posterior probabilities for genetic colocalisation=0.91, 0.84 and 0.85) (**Supplementary Table 14**). There was no association with gene expression in skin (p_FDR_ = 0.99) or the skin TWAS (p = 0.19). As CEP85 was not detectable in the mass-spectrometry proteomic analysis, we performed a second analysis using Luminex profiling of TREATGermany patients. There was no evidence for an association between EASI and serum CEP85 in this analysis (p=0.49, **Supplementary Figure 4**). However, this was not unexpected as CEP85 is not a known secreted or soluble protein.

**Figure 5.**
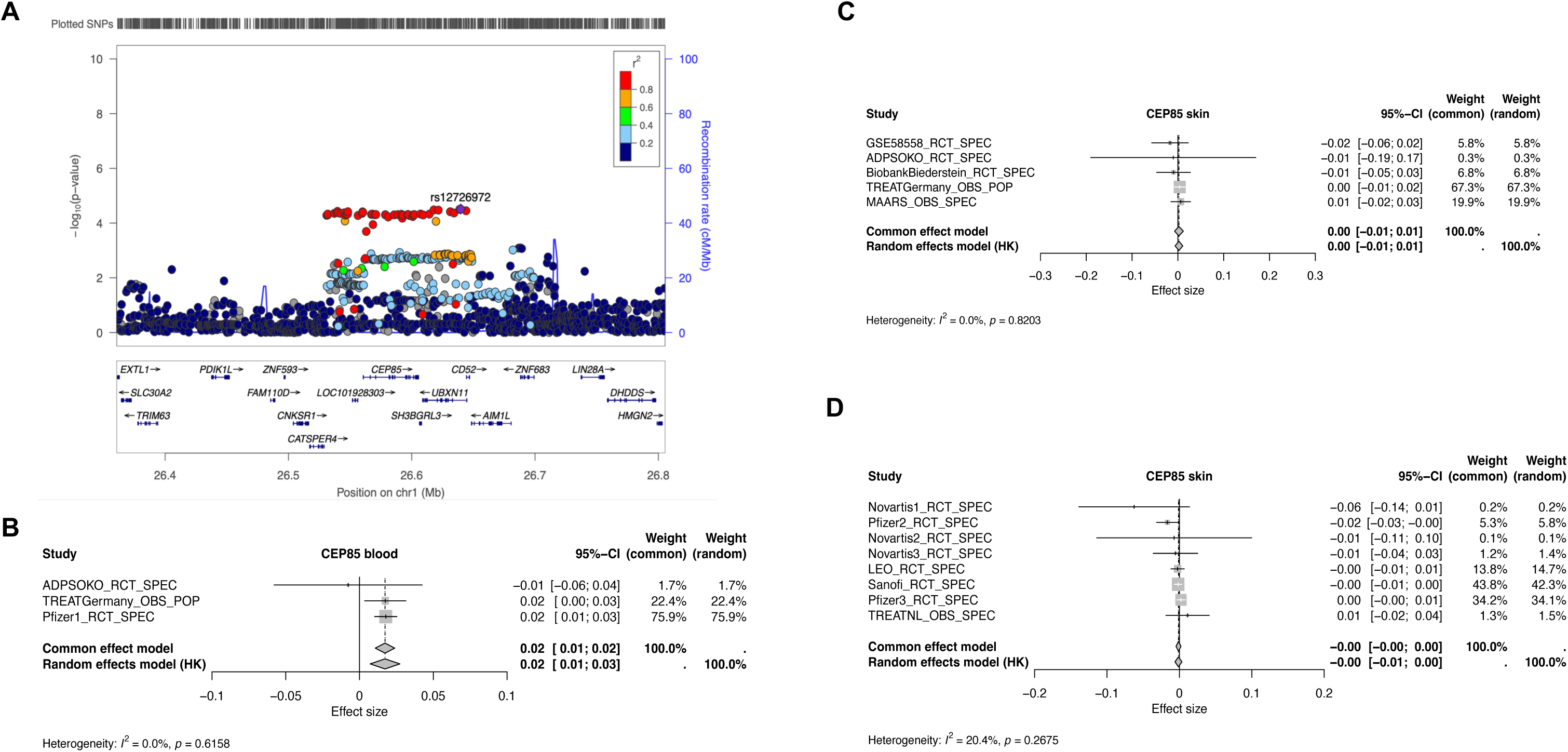
Evidence for *CEP85* association with AD severity. (A) GWAS locus zoom; (B) association with blood transcriptomics; (C) association with skin transcriptomics; (D) independent replication of skin transcriptomic association. For GWAS meta-analysis of 5 cohorts, top SNP=rs12726972, p=3.1×10^-5^, whilst didn’t reach GW significance, the TWAS result (which incorporated blood eQTL data) was significant, p=8.3×10^-13^, CEP85 was not detected in serum proteomics.

#### IL18RAP

*IL18RAP* was negatively associated with AD severity in both gene expression (beta = - 0.019, SE =0.005, p_FDR_ = 0.01) and the independent TWAS (Z-score = −8.9, p = 2.6×10^-11^) blood analyses (**Figure 6**, **Table 1**). The data did not provide evidence for an association in skin. In the blood TWAS, the *IL18RAP* signal was not independent of a nearby signal for *IL1RL1* (*R*^2^ = 0.6) as 79% of *IL18RAP* variants are also eQTLs for *IL1RL1*. However, *IL1RL1* was not correlated with EASI in the transcriptomics analysis (p_FDR_ = 0.10), suggesting that it is *IL18RAP* driving the observed signal. In addition to the genetic colocalisation between the AD severity signal and *IL18RAP* eQTL signal in blood (GTex, posterior probability of genetic colocalisation (PP.H4)=89%, **Supplementary Table 14**), strong genetic colocalisation is seen for *IL18RAP* eQTL signals for AD relevant cell types, including; CD4 T-cells^37^ PP.H4=90%, CD8 T-cells^37^=79%, CD4+_Th17 cells^38^=82% (**Supplementary Table 14**).

**Figure 6.**
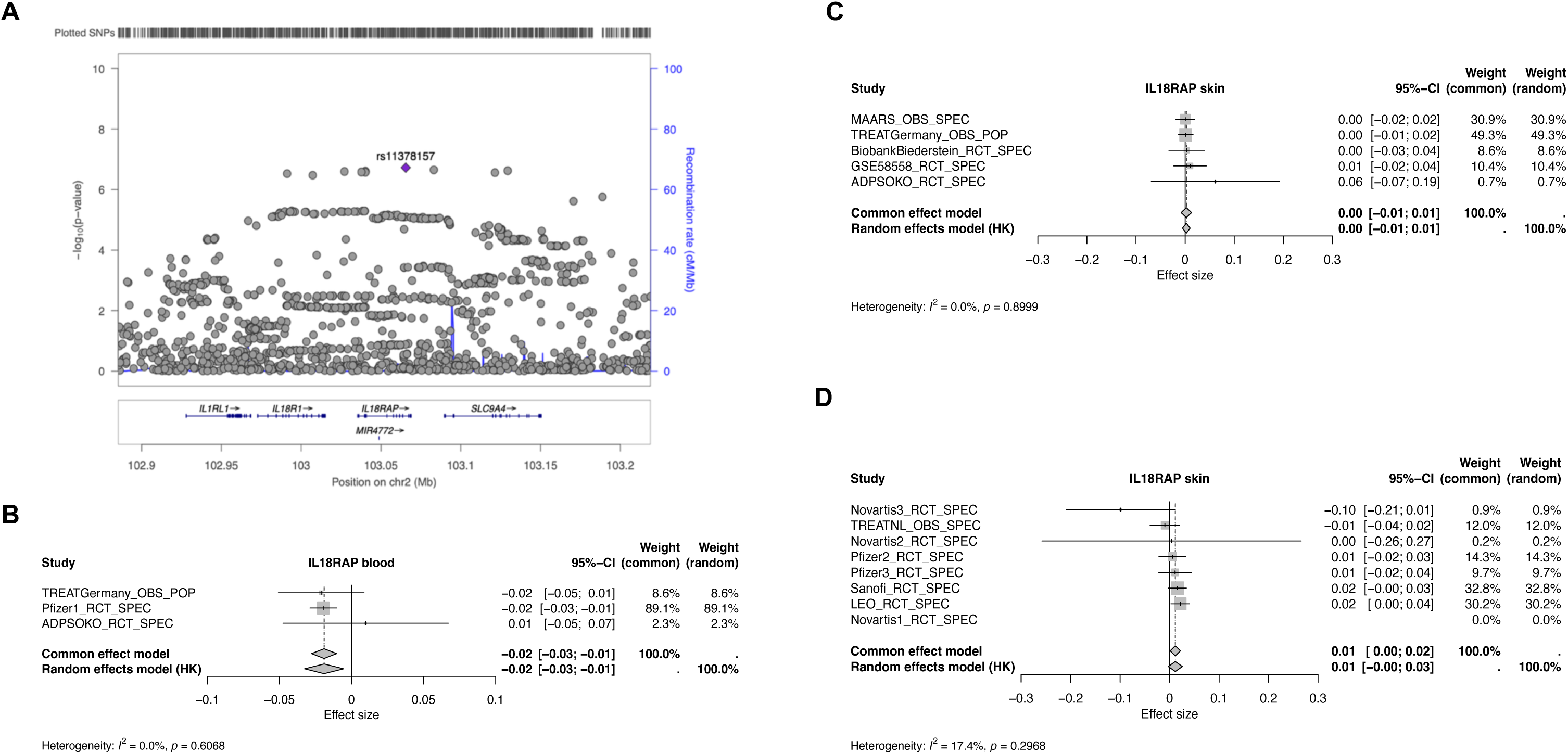
Evidence for *IL18RAP* association with AD severity. (A) GWAS locus zoom; (B) association with blood transcriptomics; (C) association with skin transcriptomics; (D) independent replication of skin transcriptomic association. For GWAS meta-analysis of 5 cohorts, top SNP=rs11378157, p=1.9×10^-7^, whilst didn’t reach GW significance, the TWAS result (which incorporated blood eQTL data) was significant, p=2.6×10^-11^, IL18RAP was not detected in serum proteomics.

IL18RAP was not detectable in the mass-spectrometry proteomic analysis, and no evidence for an association between EASI and serum IL18RAP in the Luminex profiling of TREATGermany patients (p=0.99, **Supplementary Figure 4**).

As *IL18RAP* encodes an accessory subunit of the IL-18 receptor, we investigated IL-18 and related genes/proteins. IL-18 gene expression or protein abundance was not associated with EASI in our data and there was only a weak association with *IL18R1* gene expression in blood (p=0.01, **Supplementary Table 15**).

#### PTGDR and TRIM26

*PTGDR* and *TRIM26* were both negatively associated with severity in TWAS and gene expression analysis in blood (**Table 1, Supplementary Figs 5,6**). However, we were unable to further refine these results, as there was little evidence that eQTL signal for these genes genetically colocalise with the AD severity GWAS signal in the cell types tested (**Supplementary Table 14**). Neither were detectable in the mass-spectrometry proteomic analysis, and Luminex profiling for PTGDR also failed. For TRIM26, there no evidence for an association with EASI in serum (p=0.77, **Supplementary Figure 4**). In the blood TWAS, *TRIM26* was also not independent of a signal for *RPP21* (R^2^ = 0.56). However, *RPP21* expression in blood was not associated with EASI score in our gene expression analysis (*p_FDR_* =0.73).

There was some evidence of differential expression in skin for both *PTGDR* (p = 0.002, p_FDR_ = 0.99) and *TRIM26* (p = 0.026, p_FDR_ = 0.99), at a nominal threshold although not after FDR correction.

### Replication in independent blood and skin datasets

We further tested our 4 candidate genes in a meta-analysis of 8 additional skin expression datasets (**Supplementary Table 16**) with EASI score. No relevant RNAseq whole blood datasets were available for replication. In line with the initial skin meta-analysis, none of our 4 candidates were associated with EASI at p < 0.05, under a random effects model. (**Figures 5 and 6, Supplementary Figures 5 and 6**). However, for *IL18RAP* there was a nominally significant signal in one of the cohorts (LEO Pharma data, beta = 0.019, p = 0.005), so in some cases *IL18RAP* skin expression may correlate with severity.

In the same skin expression datasets and 2 serum proteomics datasets (**Supplementary Table 16**), we also tested for replication of the 184 genes/proteins that showed significant associations in either the transcriptomic or proteomic analyses (**Fig 1**, n.b. excluding those only from TWAS, as this represents only indirect evidence). In the skin expression data, 13 genes had a p < 0.05, although none were significant after correcting for multiple testing (p<3.0×10^-4^, data received for only 169/184, **Supplementary Table 17**). In the proteomic meta-analysis of 2 datasets (n = 309), data were available for only 15 of the 184. Of the measured proteins, none were significantly associated with EASI after correction for multiple testing (p < 0.003), although five proteins (TNC, LCP1, MARCKSL1, PTK7, WARS1) were associated at P<0.05.

### CEP85 expression impacts macrophage phagocytosis

As *CEP85* has not previously been associated with AD, we undertook in vitro experiments to investigate the functional effect of *CEP85* over-expression. Our eQTL analyses identified monocytes as an important cell type for *CEP85*. Monocytes are important precursors of macrophages which are recruited into sites of skin inflammation in AD, where they undergo essential functions including pathogen clearance by phagocytosis. Although macrophages are known to accumulate in inflamed AD skin^39,40^, previous publications have suggested that phagocytosis is impaired in AD^41,42^. Using primary human monocyte-derived macrophages differentiated by M-CSF, we demonstrated a statistically significant reduction (mean difference = 5.7, 95% CI = 2.6-8.9, p = 0.006) in phagocytosis of the K562 cell line in *CEP85* overexpressing macrophages compared to control macrophages (**Figure 7, Supplementary Figure 7**).

**Figure 7.**
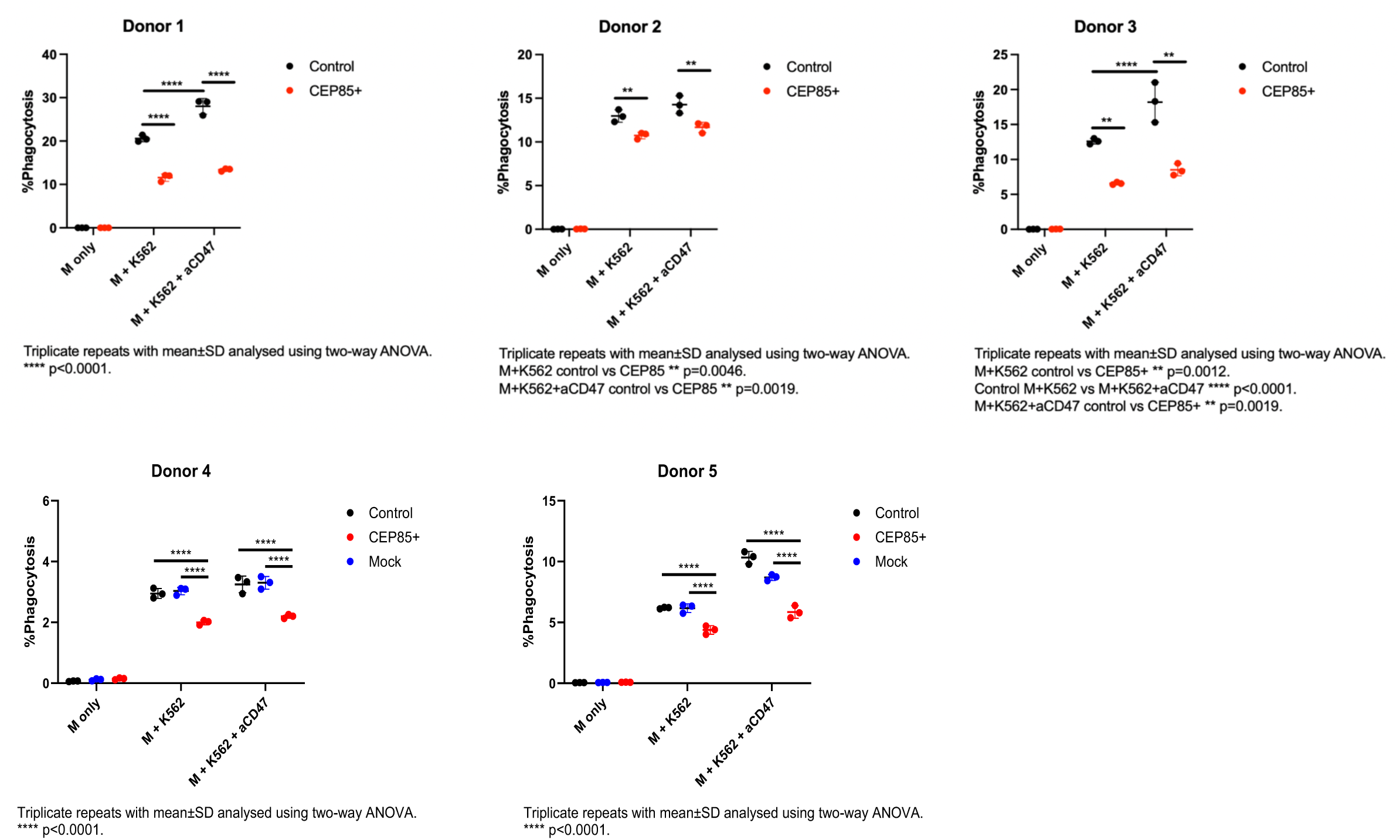
Functional effect of *CEP85* over expression on phagocytosis. Data is shown for 5 donors assayed in triplicate. *CEP85* over expressing macrophages were compared with non-transfected macrophages under three conditions. For donors 4 & 5, a mock transfection control (lipofectamine only) was also included. M only – negative control; M + K562 - cells were cultured with K652 which allows detection of phagocytosis; M + K562 + CD47 - positive control as CD47 blockade induces phagocytosis. The difference in % of cells phagocytosed was analysed using two-way ANOVA and groups compared using Fishers LSD.

## Discussion

Moderate and severe AD is associated with substantial morbidity (both directly and via systemic co-morbidities) due to the extent and degree of skin inflammation, consequent symptomatology (itch, infection, sleep deprivation) as well as comorbidities (atopic, mental health disorders, cardio-metabolic disease and treatment related sequelae such as steroid related osteoporosis^43^. Here we have used a large scale multi-omic approach to interrogate the underpinning molecular mechanisms as a path to drug development/ and predictive tools. We identify 441 genetic loci, genes or proteins associated with AD severity. We subsequently combined these results, in a triangulation approach, to identify candidates with the most robust evidence and identified 4 genes with evidence from two independent approaches.

We identify robust evidence for a role of *CEP85* in AD severity. This gene is a novel candidate for AD. GWAS has linked the gene to asthma^44^ and we found moderate correlation (R^2^ = 0.6) between the reported asthma lead SNP and our lead SNP rs12726972, suggesting it is potentially the same locus. Previous studies have found rs12726972 to be associated with monocyte^45^ and eosinophil counts^46^, immune cell types that are important in AD. The association with monocytes is particularly interesting as we found strong evidence of genetic colocalisation between the AD severity signal and *CEP85* expression in 3 separate monocyte datasets. Methylation of *CEP85* has also been associated with inflammatory bowel disease risk^47^ a comorbidity known to be also associated with AD. Current knowledge of the functional biology of *CEP85* is limited but has been shown to limit cell division by regulating the separation of centrosomes^48^. In functional in vitro follow-up of this gene, we have shown that over expression of *CEP85* in macrophages can impact cell motility by disturbing phagocytosis. It is well known that bacteria such as *Staphylococcus aureus* commonly colonise the skin of AD patients^49,50^ particularly in those with severe disease. Therefore, we hypothesise that disruption of phagocytosis of bacteria could be the mechanism by which over expression of *CEP85* contributes to severity of AD.

*IL18RAP* encodes the interleukin 18 receptor accessory protein. IL-18 is a proinflammatory cytokine that has a widely published role in AD susceptibility and some prior evidence of involvement in severity^11,22,51^, as well as other atopic and autoimmune diseases^52–55^. Whilst most published evidence suggests higher IL-18 is detrimental to disease, some studies have reported an opposite direction of effect and suggested that IL-18 may increase or decrease disease risk contingent on the cytokine environment^56,57^. The evidence around IL-18 has led to it being investigated as a therapeutic target for AD, with a phase II trial of an anti-IL-18 monoclonal antibody to treat moderate-severe AD in progress (GSK1070806, NCT04975438) and another completed (CMK389, NCT04836858). CMK389 showed signs of therapeutic efficacy, strengthening the view that this pathway is involved in AD phenotypes.

IL18RAP forms a protein complex with IL18R1 enhancing the binding of IL-18 to the receptor complex. While the IL18R1 subunit can bind to IL-18 alone, it only demonstrates a weak affinity for IL-18 binding. In contrast, the complex with IL18RAP demonstrates high affinity binding, showing the importance of IL18RAP for normal IL-18 function. In this study, we found downregulation of *IL18RAP* in blood to be associated with higher severity, consistent with studies of AD patients compared to healthy controls^56,58^. *IL18RAP* was not detected in proteomic analysis of serum, which is expected given that it is membrane-bound. The *IL18RAP* locus has been previously associated with AD susceptibility and other allergic diseases in GWAS^31,59–61^, but we show for the first time its importance in severity of the disease. In our exploration of which cell type may be particularly relevant, we found genetic colocalisation evidence with the AD severity signal to be highest in CD4+ T cells.

*PTGDR* has primarily been studied in the context of asthma^62^ but there is evidence suggesting the potential involvement of PTGDR in skin conditions including atopic dermatitis. *PTGDR* is a receptor (DP1) that mediates the effects of prostaglandin D2 (PGD2), a lipid mediator widely implicated in inflammation and allergic responses^63,64^. It is one of two receptors for PGD2 which bind with similar affinities, however we found that only *PTGDR* was associated with severity. The two receptors have distinct but complementary roles. DP1-dependent signalling has been demonstrated to promote eosinophil survival through the upregulation of pro-inflammatory genes and to mediate the vascular effect of PGD2^65,66^. The effect of PGD2 on inflammation is complex, both promoting and reducing inflammation depending on context^67^. We observed that *PTGDR* downregulation is associated with an increase in severity. Studies have demonstrated that PGD2 increases after scratching or tape stripping in patients with AD and that PGD2 inhibits pruritus and accelerates the repair of the cutaneous barrier in mice^68,69^.

*TRIM26* lies in the HLA region and has been associated with asthma^70^ but not any other atopic disease. However, studies have found TRIM26 to be involved in immune^71,72^ and inflammatory responses^73^, similar to other *TRIM* family genes. A murine study demonstrated *TRIM6* is a positive regulator of toll-like-receptor, TNF-α-, and IL-1β-induced inflammatory responses through regulation of TAK1 activation^73^. All of these pathways have been associated with AD, although primarily in skin (through barrier function) rather than blood^74,75^.

We have conducted analyses in both blood and skin tissues. Whilst more results were found for blood, there are important factors to consider as to why we do not see signals in skin. There was a substantial difference in power (due to sample size) of the TWAS analyses (blood eQTL data sample size is 85x larger than skin data) and the gene expression analysis (blood meta-analysis sample size was 1.8x larger than for skin). EASI score is also a measure of total skin disease involvement and inflammation, and so blood may better represent this burden of inflammation. Additionally, skin samples are likely to be more heterogeneous (e.g. due to variability in skin composition related to biopsy site and/or degree of subclinical disease activity), making signals harder to detect. We also found evidence that the surrogate variables, used to control for unmeasured variability such as batch effects, correlate with severity (**Supplementary Figure 2**), and hence the analysis may attenuate real signal.

There are various other limitations to our work. While we included an analysis of protein abundance from one study in blood serum, this was relatively small (N=75) and measured only ∼2,000 proteins, limiting the triangulation potential for this data.

It is also important to note the heterogeneity of severity definitions across our analyses. Our gene expression and protein abundance analyses measured disease activity at a single timepoint, which may not indicate severity over time. In addition, these cohorts were predominantly from dermatology specialist centres and are not representative of the full spectrum of AD disease severity in the population, hence could be affected by referral bias (towards more severe disease). In contrast, while validated objective disease severity scores are not available in large population cohorts (as used in the GWAS) a more longitudinal history can be captured, but only with a proxy severity measure (hospitalisation and use of systemic and potent therapies that will only be prescribed for those with the most severe AD). This measure is likely to vary between health systems (and by the multiplicity of factors influencing healthcare use) but permits analysis of larger numbers of patients (N>100,000 in the GWAS). In addition, we recognise the limitation that studying AD severity in existing large-scale cohorts and trials restricts us to a narrow definition that excludes wider disease severity concepts important to individuals, such as comorbidities and well-being^7,76–78^. We also acknowledge the lack of representation from non-European populations in our analyses. Future data collection should focus on under-represented populations to ensure that studies can address this gap and ensure that health inequalities are reduced rather than exacerbated^79^.

Whilst we consider it a strength that we bring together multiple omics (and study designs) in this analysis, there are multiple reasons why a gene/protein may not be associated in all analyses. Different omics layers represent different aspects of the biological pathway and so an association seen at the gene level is not always expected to be seen at the protein level, for example. Clearly there will be differences between effects seen in different tissues, and we also have included different ways of measuring severity, which whilst correlated represent slightly different aspects of the phenotype. Additionally, our approach to integrate genetics into the analysis identifies genes that are likely to be causally related to AD severity, rather than just being a biomarker for disease activity. Therefore, for the 4 genes where we do see consistency across analyses that have used both EASI score and proxy measures from electronic health records, as well as being implicated by both genetic and gene expression evidence, these genes may have a broad causal impact on AD severity. However, there are likely to be important findings amongst our results from each analysis, that we have not focused on here, so future studies are warranted to investigate many of these further. We therefore share the full results to enable further investigations of these additional candidate genes and proteins.

For the molecular markers we have observed to be associated with disease severity, we incorporated genetic evidence to investigate causality. Therefore, the genes identified may be particularly relevant as drug targets for severe AD. In addition, they may also serve as valuable biomarkers for prediction of (and potentially monitoring for) severe disease. However, to achieve these goals, further work is necessary to examine these molecular candidates in longitudinal studies for prediction and monitoring, combined with experimental studies to evaluate potential drug targets. In the context of biomarker discovery, numerous ethical and social considerations are required for their use in a clinical setting^80,81^.

In this large-scale study we have highlighted 4 genes (*CEP85*, *IL18RAP*, *PTGDR*, *TRIM26*) for which there is multi-omic evidence for involvement in AD severity and provide support that these associations are causal. Together this work represents a strong basis from which to examine the role of each candidate protein in risk prediction, for disease monitoring and as novel drug targets, to provide much-needed advances in clinical care. In particular we note that *CEP85* is a novel candidate for AD severity for which we have found genetic and transcriptomic evidence, and we demonstrate what the functional mechanism may be with in vitro work.

## Methods

All statistical tests are two-tailed unless otherwise specified.

### GWAS

#### GWAS phenotype definition

Within individual cohorts, participants with AD were identified. Criteria for defining AD included “ever had atopic dermatitis” according to doctor-diagnosed or self-reported data.

Individuals with AD were then categorised as either having severe AD or not. The following criteria were used; use of systemic medication to treat AD (**Supplementary Table 18**), use of potent/super potent topical medication to treat AD on two or more occasions (**Supplementary Table 18**), hospitalisation with AD as the primary diagnosis (**Supplementary Table 19**), visited a dermatology specialist, or ever had phototherapy treatment. Those that had at least one of the qualifying criteria were classified as having severe AD and all others, without any missing data for the fields used to define the phenotype, were classified as non-severe. Specific cohort definitions for severity can be found in Supplementary methods.

For the GWAS, we compared individuals with severe AD (cases) to those who had AD but did not meet the criteria for severe disease (controls).

#### GWAS and Quality Control

All cohorts are European and were imputed individually either using the haplotype reference consortium reference panel^82^ or to a population specific panel. Full details of genotyping, imputation, quality control (QC) and GWAS analysis for individual cohorts can be found in supplementary methods. All cohorts performed a GWAS under a logistic model using age, sex and relevant ancestry PCAs as covariates. All cohort summary statistics also underwent additional quality control prior to meta-analysis, to standardise QC. Variants with MAF < 1% or imputation scores less than <0.7 were removed.

#### GWAS meta-analysis

We performed a GWAS meta-analysis for AD severity across 5 cohorts including 25,744 cases defined as severe and 75,022 defined as non-severe (controls) of European ancestry. The meta-analysis was performed using GWAMA^83^ 2.2.2 under a fixed effects model. Approximately 11 million variants in total were included in the meta-analysis.

#### Gene prioritisation

##### Fine-mapping

Fine-mapping was performed for the chromosome 11 locus. Conditional regression was first used to identify the number of causal variants and fine-mapping was then performed using PAINTOR^84^. Credible sets of causal SNPs were then assembled for 95% coverage.

##### Colocalisation

For the 4 genes of interest, we performed genetic colocalization between the GWAS and eQTL signals (for various tissue and cell types) using the coloc method in R. We downloaded summary statistics for skin, whole blood and immune cell types from the eQTL Catalogue^85^ and Open GWAS^86^. We considered a PP.H4 >80% to be strong evidence of a shared causal variant.

##### Mendelian randomisation

Using the GWAS results and blood eQTL data from eQTL Gen^87^ (31,684 samples), we performed mendelian randomisation (MR) analyses of severity for the 4 genes of interest. MR analyses were performed using the TwoSampleMR package in R. The eQTL summary statistic data was downloaded from OpenGWAS for the 4 genes of interest. MR estimates were generated using the inverse variance weighted (IVW) method. Both cis and trans-eQTLs were considered as instruments. F-statistics were generated for each instrument, to confirm relevance assumption.

### TWAS

TWAS for blood and skin were performed using the software OTTERS^88^. For the blood TWAS cis-eQTL summary statistic data from eQTLGen was used and for the skin TWAS, cis-eqtl summary statistic data from TwinsUK-skin was used. For both TWASs, the European panel of 1000 genomes was used as the reference panel for calculating LD in stage 1 and the AD severity GWAS summary statistics generated in this study used in stage 2.

To reduce test-statistic inflation, outputted summary statistics were corrected using the *bacon* Bioconductor package in R. As outlined in the OTTERS protocol, we confirmed the median p-value was equal to 0.5 after this adjustment. The number of independent genes was identified by calculating squared Pearson’s correlation between every pair of significant genes. Gene pairs with R^2^ < 0.5 were considered to be independent of each other.

### Differential gene expression (DGE) analyses

#### Blood

Transcriptomic profiles based on 340 AD blood samples have been acquired from academic (P2N_ADPSOKO, TREATGermany) or industrial sources (Pfizer1). Blood samples have been profiled using RNAseq technology (P2N_ADPSOKO, Illumina NovaSeq 6000/HiSeq 2500; TREATGermany, Illumina HiSeq4000; Pfizer, Illumina NovaSeq). In all cohorts the samples and severity scores used in this analysis were collected/measured at a baseline timepoint in individuals who were not undergoing systemic treatment. Further cohort details can be found in Supplementary methods.

#### Skin

Transcriptomic profiles based on 185 AD non-lesional biopsies have been acquired from public (MAARS, GSE58558) or academic sources (BiobankBiederstein, P2N_ADPSOKO, TREATGermany). Punch biopsies have been profiled using microarray (MAARS, Affymetrix Human Gene 2.1 ST Array; GSE130588, Affymetrix Human Genome U133 Plus 2.0 Array) or RNAseq technology (BiobankBiederstein, Illumina HiSeq4000; P2N_ADPSOKO, Illumina NovaSeq 6000/HiSeq 2500; TREATGermany, Illumina HiSeq4000). In all cohorts the samples and severity scores used in this analysis were collected/measured at a baseline timepoint in individuals who were not undergoing systemic treatment. Further cohort details can be found in Supplementary methods.

#### Raw data processing

Raw microarray data (Affymetrix CEL files) has been processed using single-channel array normalization (SCAN) employing the R package SCAN.UPC version 2.30.0^89^. Raw sequencing data has been processed using TrimGalore version 0.6.6, Cutadapt version 4.4, FastQC version 0.11.9, HISAT2 version 2.2.1^90^, SAMtools version 1.11^91^ and Rsubread version 2.2.6^92^ and annotated according to Ensembl release 103 (GRCh38). Sequencing count data has been converted to the logarithmic scale using the voom transformation provided with R package limma version 3.60.6^93^ and scaling factors estimated using R package edgeR version 4.2.2^94^. Harmonization of molecular and clinical data (sex, age, (objective) SCORAD, EASI) incorporated in the meta-analyses has been performed using the R package harmonizeGeneExprData, available at GitHub (https://github.com/szymczak-lab/harmonizeGeneExprData). Subsequent quality control of the resulting datasets has been performed using the R Package QCnormSE, available at GitHub (https://github.com/szymczak-lab/QCnormSE).

For all studies, expression values of each gene have been standardized to a mean of 0 and standard deviation of 1 to increase comparability across profiling technologies.

#### Measuring disease severity

Disease severity has been measured in terms of predicted EASI (pEASI) as implemented in R package mapSeverityScores^8^ derived from objective SCORAD or SCORAD (in this order, dependent on data availability).

#### Differential expression analysis

Single-cohort DGE has been performed using linear model

expr∼pEASI+sex+age+pEASI:sex+pEASI:age+SV. Surrogate variables (SV) have been estimated using R package sva version 3.52.0, and regression coefficients have been estimated using R package limma version 3.60.6. Random effects meta-analysis for estimating pooled regression coefficients (corresponding to (log2) FC) has been performed using the R package metaAnalyzeGeneExprData (https://github.com/szymczak-lab/metaAnalyzeGeneExprData), providing wrapper functions for R package metafor version 4.6-0^95^. Resulting meta-analysis p-values were adjusted for multiple testing using the Benjamini-Hochberg procedure and significantly differentially expressed genes were required to meet adjusted p-value < 0.05. Visual and tabular summaries of analysis results have been generated using R packages tidyr version 1.3.1 and sjPlot version 2.8.16.

### Differential protein abundance (DPA) analysis

Serum samples were depleted of abundant proteins using the Human-14 Multi Affinity Column (Agilent) before data-independent acquisition (DIA) LC-MS and data-dependent acquisition (DDA) MS/MS data collection. Mass spectrometry spectral libraries were generated using a modified TOP15 method^96^. DIA mass spectrometric data were analysed with Spectronaut software (Biognosys, version 15.01).

A total of 2217 proteins were quantified. The protein intensities were log-transformed, batch corrected, and quantile normalized before statistical modelling analysis, with the LC column included as a covariate. p-value correction for multiple testing was performed using the Benjamini-Hochberg method to control the false discovery rate (FDR). All analyses were conducted using R version 4.0.0.

Candidate proteins were also measured in 374 baseline serum samples from TreatGermany using Luminex. Carbodiimide coupling of the capture antibody (CCDC21 polyclonal antibody, Proteintech, 26314-1-AP) to MagPlex®-Microspheres as well as subsequent assessment of coupling efficiency was performed according to the protocols described in the xMap® Cookbook (Edition 5). Subsequent sample analysis was performed according to the protocol “Capture Sandwich Immunoassay” described in the xMap® Cookbook ((Edition 5) with the following minor adaptations: PBS-TBN buffer was used as wash buffer. Microspheres were incubated with 10 µl of sample or standard (Recombinant protein of human coiled-coil domain containing 21 (CCDC21), Origene, TP309507) in a total assay volume of 100 µl for 2 hours at room temperature. Biotinylated detection antibody (CEP85 polyclonal antibody, Thermo Fisher Scientific, A303-326A) was diluted to a final concentration of 1,5 µg/mL and incubated with the microspheres for 1 hour at room temperature.

All data analysis were done in the statistical computing environment R (R Core Team 2019). Four-point logistic models were fit to the standards using package dr4pl. The relationship between protein level and EASI score was tested using linear regression.

### AD disease map

AD severity candidates were mapped to the Atopic Dermatitis disease map^32^ using HGNC identifiers. We identified candidate proteins overlapping with the contents of the map and visualized them as a dedicated data overlay to highlight mechanisms and cell types involving these candidates.

We then investigated severity candidates not matching the AD map. We used the BioKB text mining platform (https://biokb.lcsb.uni.lu/) to identify interactions between these novel candidates and proteins in the AD map and filtered these candidates using the DisGeNET database to shortlist for those linked with conditions annotated either with “skin diseases” and “inflammatory diseases”.

### Network analysis of significant biomarkers

A co-expression network was constructed using curated and pre-processed transcriptomic data from the TREATGermany, ADPSOKO, and Pfizer1 cohorts. Batch effects from the different data sources were mitigated using the pamr.batchadjust function from the pamr R package^97^. Transcript IDs were mapped to gene symbols using the bitr function from the clusterProfiler package^98^, and expression values corresponding to the same gene symbol were aggregated by taking the median. The co-expression network of blood samples was then inferred via the INfORM algorithm^99^, using CLR^100^ as the inference method and Spearman correlation as the estimator. Several network centrality measures (degree, betweenness, clustering coefficient, closeness, and eigenvector) were calculated, and a final centrality rank was compiled by aggregating individual centrality ranks via the Borda function from the TopKLists R package^101^.

The severity module was computed using the Diamond algorithm^102^, with severity biomarkers derived from the TWAS and transcriptomic experiments serving as seed genes. To further validate functional relationships, the co-expression network was filtered to retain only edges supported by experimental evidence in a protein–protein interaction (PPI) network described by Cheng et al^103^. Shortest-path analysis was repeated on this filtered network. Functional annotation of the severity module was conducted using the cReactome PA Bioconductor package.

### Triangulation

We performed triangulation on a gene-based level, aligning TWAS, transcriptomics DGE and proteomic DPA results, to identify genes significant (after multiple test correction) in two or more of the independent omic analyses. However, we did not triangulate between skin and blood TWASs as they were computed using the same GWAS results and therefore we considered them not independent.

### Replication analysis

The 184 genes / proteins that were significant in either transcriptomic or proteomic analysis were taken forward for replication. We received transcriptomic summary statistic data for our candidates from 7 randomised controlled trial (RCT) datasets: 1 from LEO Pharma, 3 from Novartis, 2 from Pfizer, 1 from Sanofi and from 1 observational dataset: TREAT_NL. Cohort descriptions and trial identifiers are provided in **Supplementary Table 16**. Cohorts performed DGE analysis using the model: Expression ∼ AD severity + age + sex + relevant batch effects (determined by the cohorts). We limited the analysis to individuals of European ancestry only. Overall, we received data for 169 / 184 candidates from at least one cohort. We meta-analysed the results for the 169 genes under a random effects model in R (v4.4.2).

We also received serum proteomic summary statistic data from 2 Novartis datasets (**Supplementary Table 16**). Novartis performed proteomic analyses using the model aptamer ∼ EASI + SEX + AGE + relevant batch effects + additional relevant covariates. We performed a similar meta-analysis as described above; however, data were available for only 15 proteins of the 184 in a total of 309 individuals of European ancestry.

### *CEP85* experimental data

#### PBMC isolation

Peripheral blood monocyte cells (PBMCs) were isolated from fresh human leukopaks (Cambridge Bioscience) by diluting 1:1 with DPBS (Gibco), layering onto Ficoll-Paque and centrifuging at 400xg, 30 mins. The PBMC interface was collected and washed in DPBS by centrifuging at 300xg, 8 mins. The isolated PBMC were frozen in 90% FBS, 10% DMSO and stored in liquid nitrogen.

#### mRNA synthesis

CEP85 over expression plasmid was purchased from Genscript using the HEK optimised DNA sequence. The DNA template for mRNA synthesis was generated via PCR using Platinum™ SuperFi™ PCR Master Mix (Invitrogen 12358010) according to the manufacturer’s protocol, with primers designed to amplify the desired DNA sequence containing the T7 promoter. A sequence of 100 adenine (A) nucleotides was introduced to 3’ end of PCR via reverse primer. PCR product was subsequently purified using QIAGEN QIAquick PCR Purification Kit. The *in vitro* transcription (IVT) of mRNA was carried out using MegaScript™ T7 Transcription Kit Plus (Invitrogen A57622) and co-transcriptionally capped using the CleanCap® reagent (TriLink Biotechnologies) following the manufacturer’s protocol and purified using the QIAGEN RNeasy Midi Kit. The integrity and purity of the RNA were assessed by measuring the A260/A280 ratio using a NanoDrop™ spectrophotometer (Thermo Fisher Scientific) and verifying RNA integrity via agarose gel electrophoresis.

#### Macrophage transfection

Monocytes were purified from PBMC donors using a pan monocyte isolation kit (Miltenyi Biotech) and cultured for 6 days in low-attachment 24-well plates in 10% FBS, 1% Glutamax, 1% penicillin/streptomycin in RPMI (all Gibco) with 50ng/mL M-CSF (Merck) to obtain monocyte-derived macrophages. Medium containing M-CSF was refreshed every 2-3 days. On day 6, macrophage medium was changed to 5% FBS, 1% Glutamax, 50ng/mL M-CSF in RPMI, and wells were transfected with 1.5uL lipofectamine messengerMAX (Invitrogen) and 1ug CEP85 mRNA in 50uL OptiMEM. Negative control wells for assessment of phagocytosis were non-transfected. EGFP mRNA (Trilink) was added to a single well as a transfection control and cultures were incubated overnight at 37°C, 5% CO2.

#### Phagocytosis assay

K562 cell line was labelled with CellTrace Yellow (Invitrogen) as the target cells. Macrophages were cultured at 2:1 with K562 cells in 10% FBS, 1% GlutaMAX, 1% penicillin/streptomycin in RPMI for 2h at 37°C, 5% CO2. Anti-CD47 antibody was used at 10ug/mL as a positive control to induce phagocytosis by blocking the CD47 “don’t eat me signal” on K562 cells. Cells were stained for flow cytometry using 10% Fc block in FACS buffer (0.5% BSA, 2mM EDTA in DPBS) for 15 mins at 4°C, followed by CD11b APC 1in100 in FACS buffer for 20 mins at 4°C, and resuspended in 500ng/mL DAPI in FACS buffer. Samples were acquired using a ZE5 flow analyser (Bio-Rad) and analysed using FlowJo software (v10.8.1).

#### CEP85 western blot

Protein was collected using RIPA buffer + 1% Halt protease/phosphatase inhibitor cocktail (Thermo Scientific). Protein concentration was determined by BCA assay (Thermo Fisher Scientific) and 20ug combined with NuPAGE LDS sample buffer (Invitrogen) and NuPAGE reducing agent (Invitrogen) before heating at 95°C for 5 mins. Samples were loaded into a Bolt 4-12% Bis-Tris Plus wedge-well gel (Invitrogen) and separated by electrophoresis at 180V for 35 minutes. Protein was transferred onto PVDF membranes using an iBlot 2 (Invitrogen), blocked for 1h in 5% milk, 0.05% Tween in DPBS and incubated overnight with 1in1000 anti-human CEP85 antibody (Proteintech, 26314-1-AP), followed by 1in1000 goat anti-rabbit-HRP secondary antibody (Agilent Technologies, P0448) for 1h. Beta-actin was measured as a loading control using beta-actin-HRP (Santa Cruz Biotechnology, sc-47778 HRP). Membrane washes were performed using 5% BSA, 0.1% Tween-20 in DPBS and antibodies were diluted in 5% BSA, 0.05% Tween-20 in DPBS. Chemiluminescence was measured using ECL Prime substrate (Thermo Scientific) and imaged using an iBright (Invitrogen). Band density was measured using ImageJ v1.53c. **Supplementary Figure 7** shows western blot results for the 5 donors.

## Data availability

Full summary statistics for all omic analyses are available at https://biomap-dataviz.lcsb.uni.lu/. Code used to perform the GWAS analyses and for performing triangulation can be found at https://github.com/Katie-Watts/AD-Severity. Code for running the transcriptomic analysis can be found at (https://github.com/szymczak-lab/).

## Acknowledgements

This project has received funding from the Innovative Medicines Initiative 2 Joint Undertaking under grant agreement number 821511 (Biomarkers in Atopic Dermatitis and Psoriasis). The Joint Undertaking receives support from the European Union’s Horizon 2020 research and innovation programme and the European Federation of Pharmaceutical Industries and Associations. KW, JM, LP work in a research unit funded by the UK Medical Research Council (MC_UU_00032/1 and MC_UU_00032/3). LP also acknowledges support from the LEO Foundation (LF-AW_EMEA-23-40027). This work was carried out using the computational facilities of the Advanced Computing Research Centre, University of Bristol—http://www.bristol.ac.uk/acrc/.

We thank Elke Rodriguez for her expert support with P2N_ADPSOKO, Madison Mack for graphic design of Figure 1, and Jing Zhang for generation of the *CEP85* mRNA. Renza for data management.

Individual cohort acknowledgements are in the Supplementary Methods.

## Author contributions

Designed and co-ordinated the study: LP, SB, CS, KW. Performed GWAS meta-analysis: KW. Performed transcriptomics meta-analysis: MH, SS, JH. Performed proteomic analysis: HX, XL, BZ, MH, KV. Designed/performed in vitro experiments: JR, HC. Performed network analysis: AF. Performed statistical analysis within cohorts: SL, TB, KY, MT-L, LT, WO, IH, EM-D, LR, HX. Atopic dermatitis map: MO, OL, MA. Data acquisition/supported analysis/interpretation of data: RR, JS, SW, AB, M-CF,SL,NL,SE, EVB, SL, TB, MT-L, LT, KY,BB, WO, MMH, CC, EDR, JK, LH, SK, KK, KT, KV, CD, IH, PL, KP, EM-D, AL-AB, LR,SB,FB, KK, JS, TW, BJ, PS-N, ML, AH, VS, JM, ND, SW, DG, KV, MO, NF, RB, SG, JR, SJB, CS, LP. Production of Data-Viz website: AK, SG, VS. Wrote first draft of manuscript: KW, LP, SB, CS. All authors reviewed the paper and provided feedback

## SUPPLEMENTARY FIGS

**Supp Figure 1.**
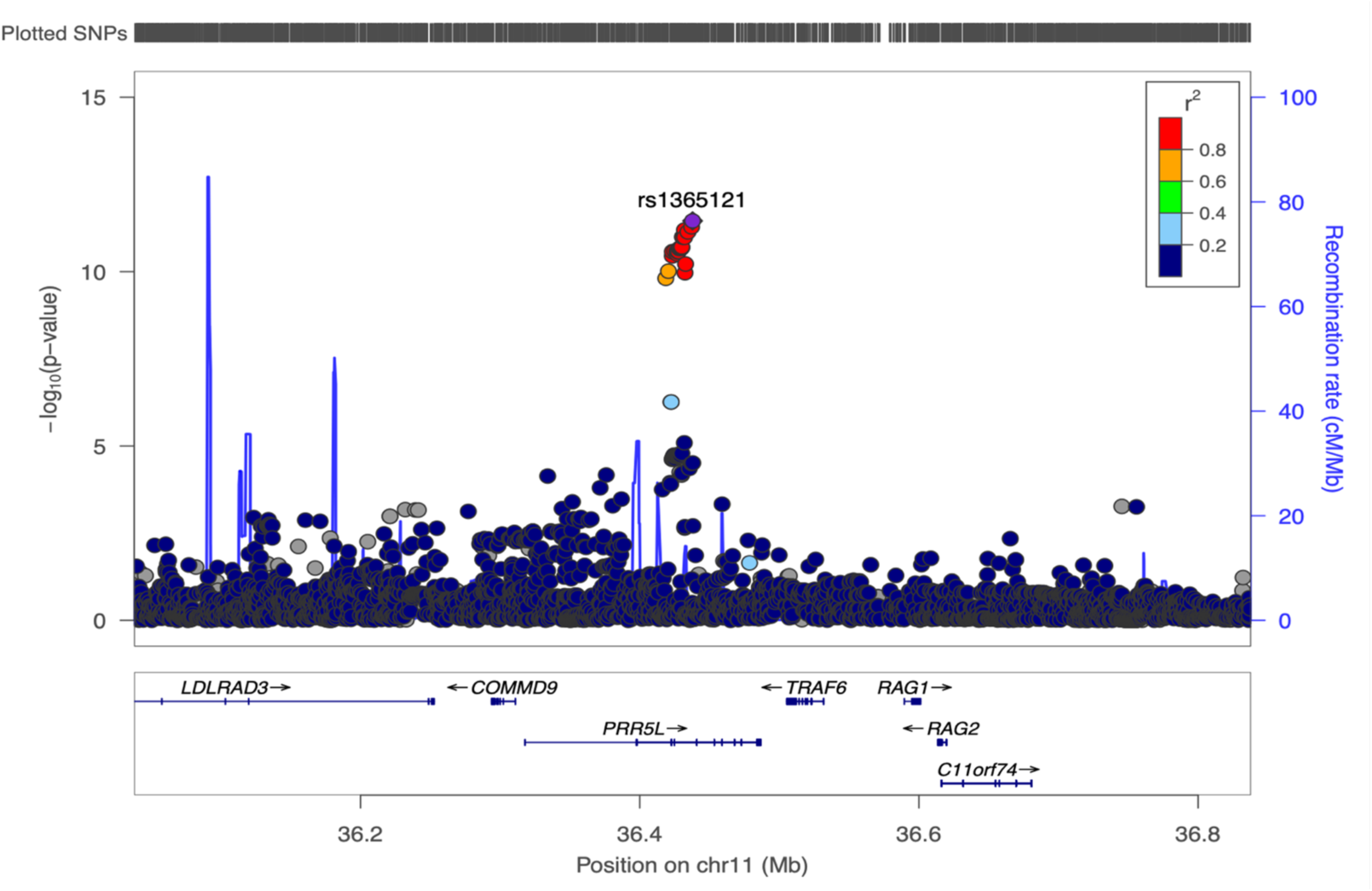
LocusZoom plot of chromosome 11 GWAS signal.

**Supp Figure 2.**
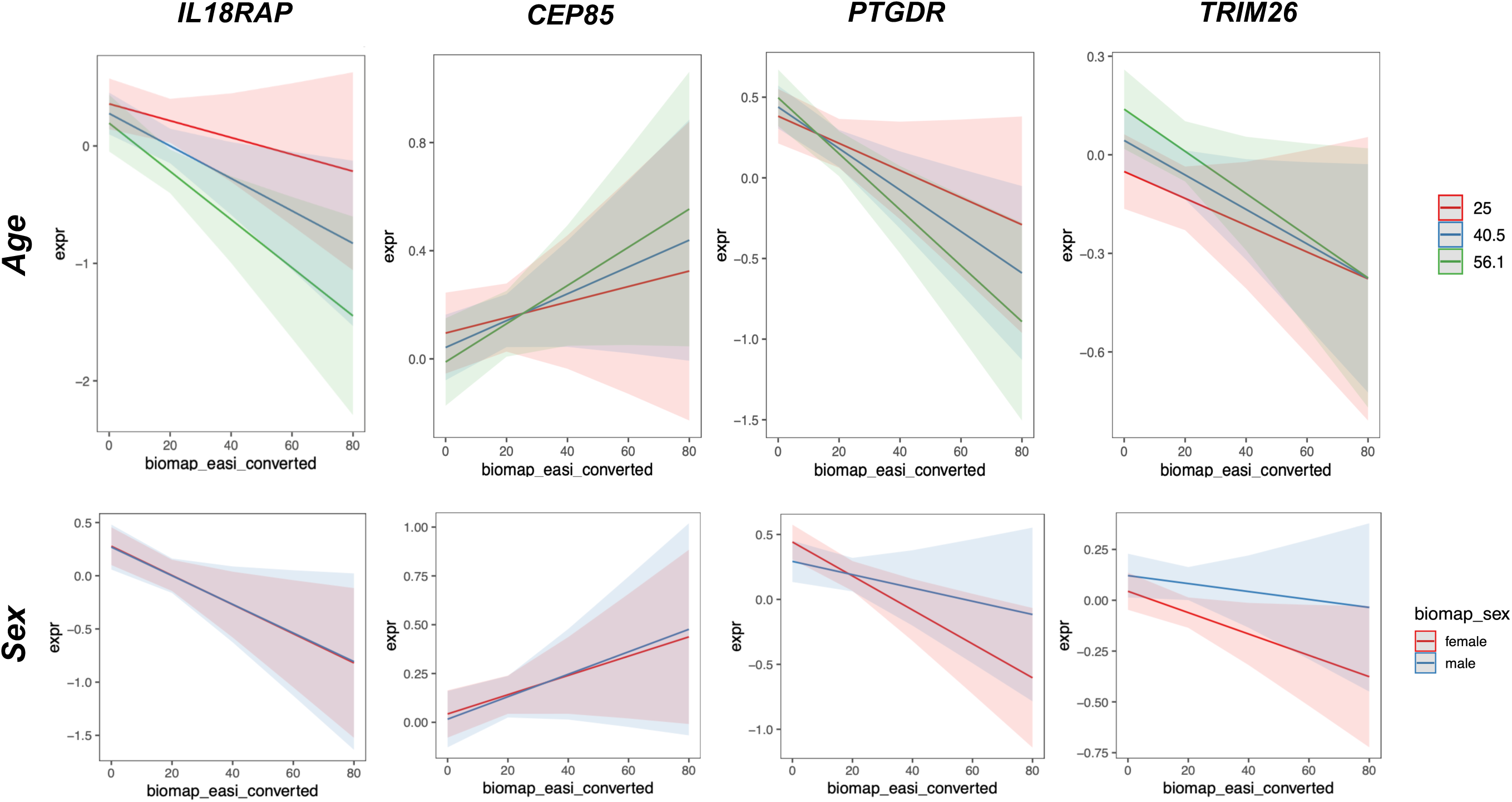
Interactions between EASI:age and EASI:sex in Pfizer1 blood cohort for the 4 genes from triangulation.

**Supp Fig 3.**
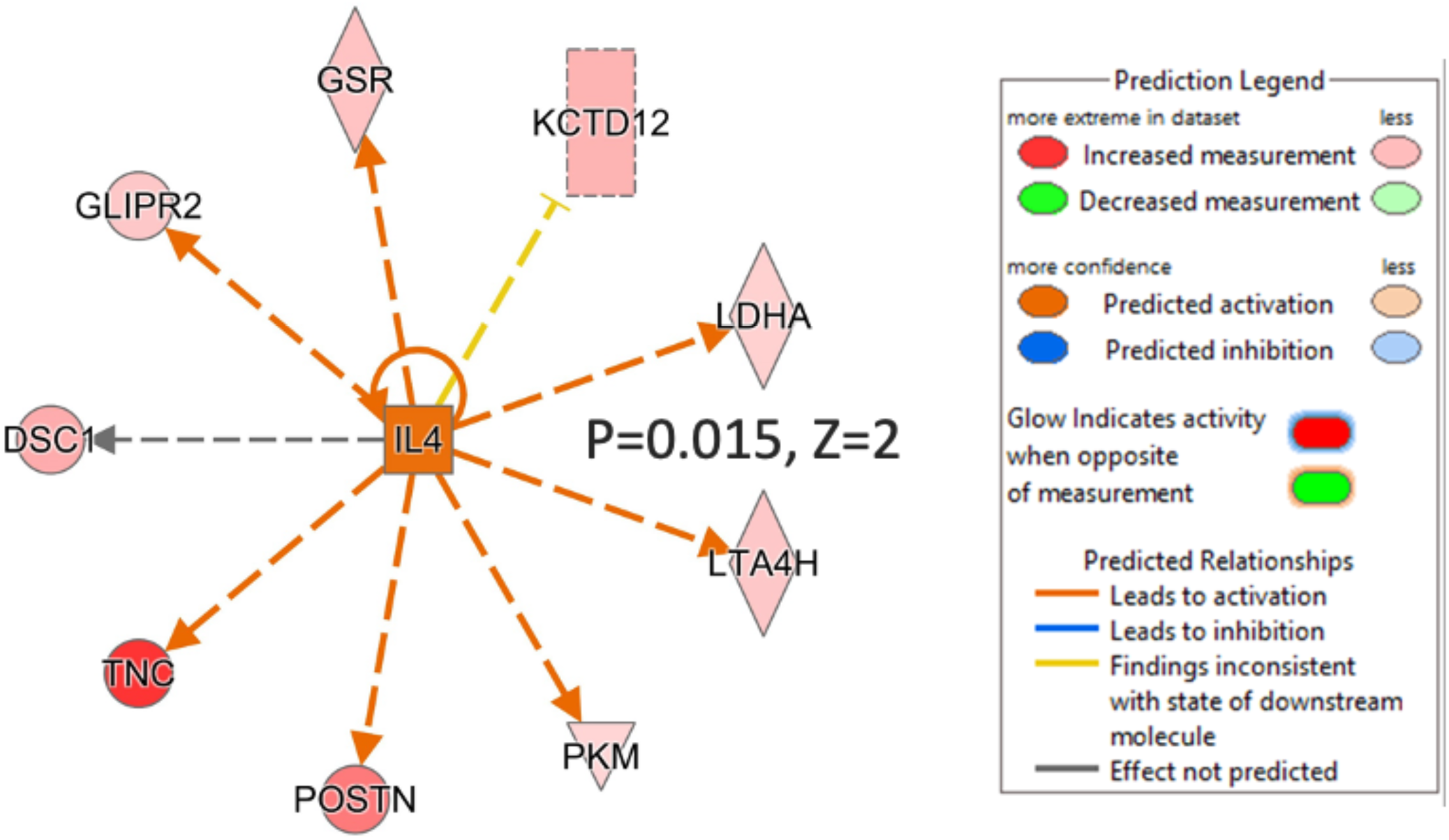
QIAGEN ingenuity pathway analysis of significant proteins highlights IL4 signalling.

**Supp Fig 4.**
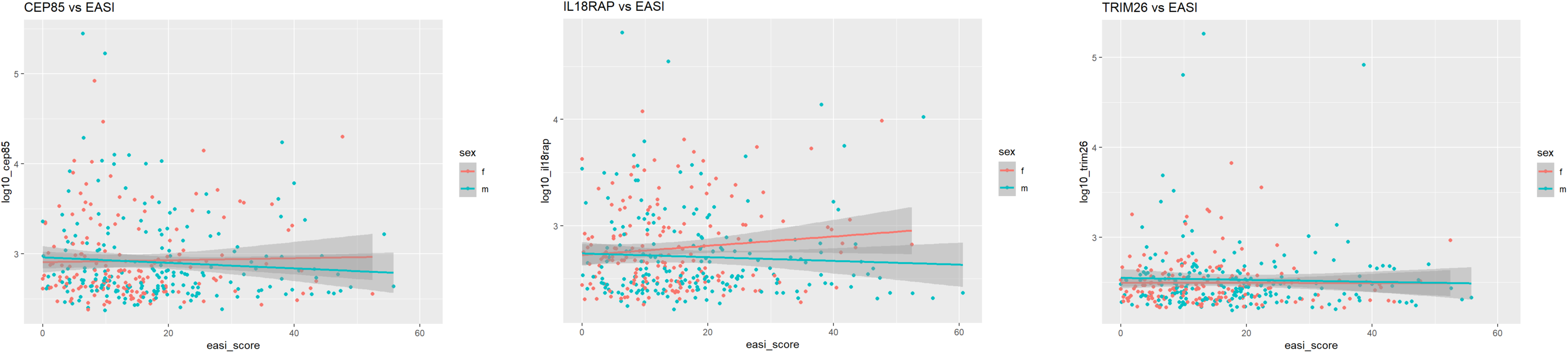
Association of EASI with serum proteins in TreatGermany. (A) *CEP85* was detectable in 357 samples. No association between serum levels and EASI score was observed (p = 0.49). (A) *IL18RAP* was detectable in 362 samples. No association between serum levels and EASI score was observed (p = 0.99). (A) *TRIM26* was detectable in 362 samples. No association between serum levels and EASI score was observed (p = 0.77).

**Supp Figure 5.**
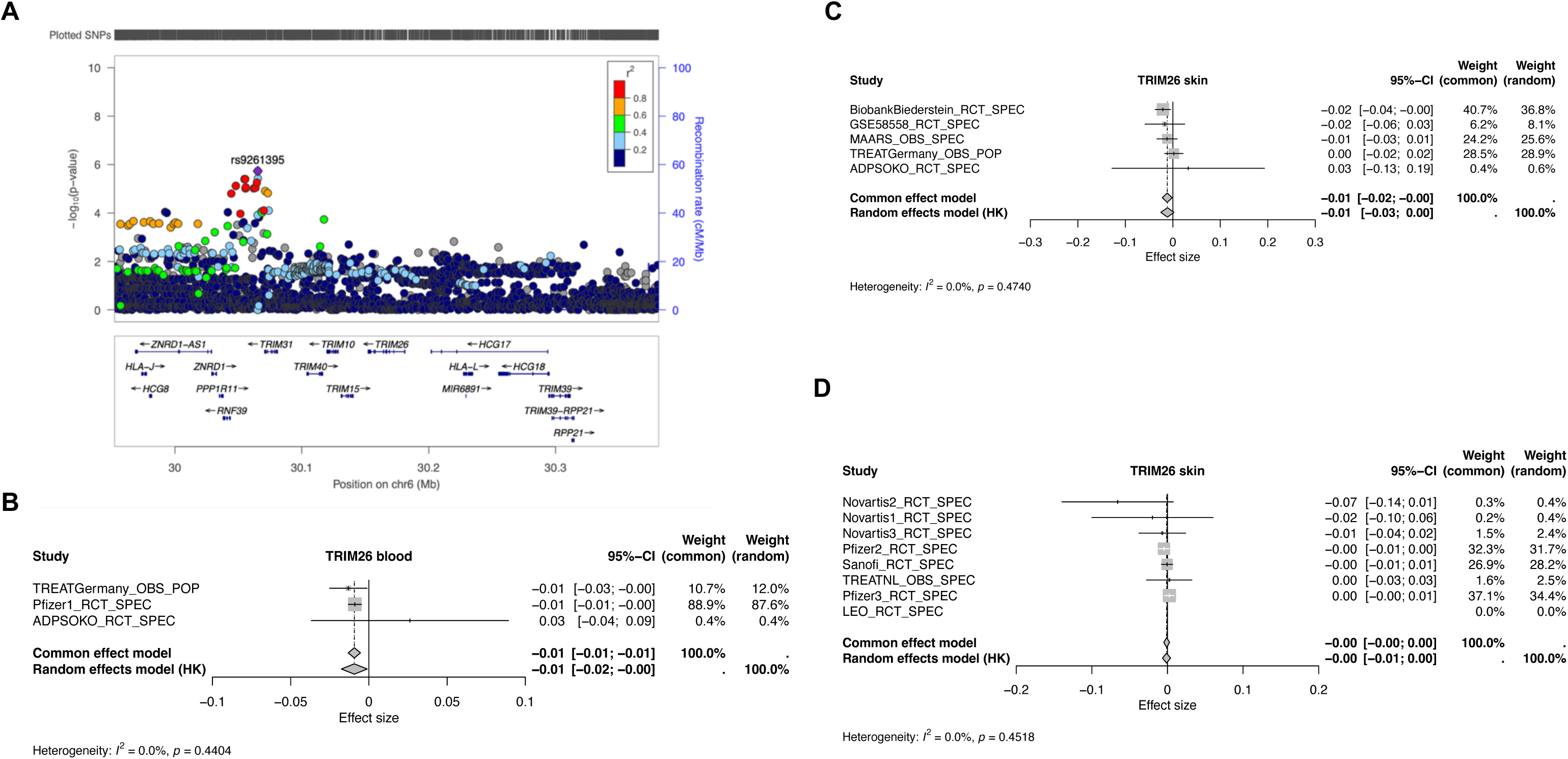
Evidence for *TRIM26* association with AD severity. (A) GWAS locus zoom; (B) association with blood transcriptomics; (C) association with skin transcriptomics; (D) independent replication of skin transcriptomic association.

**Supp Figure 6.**
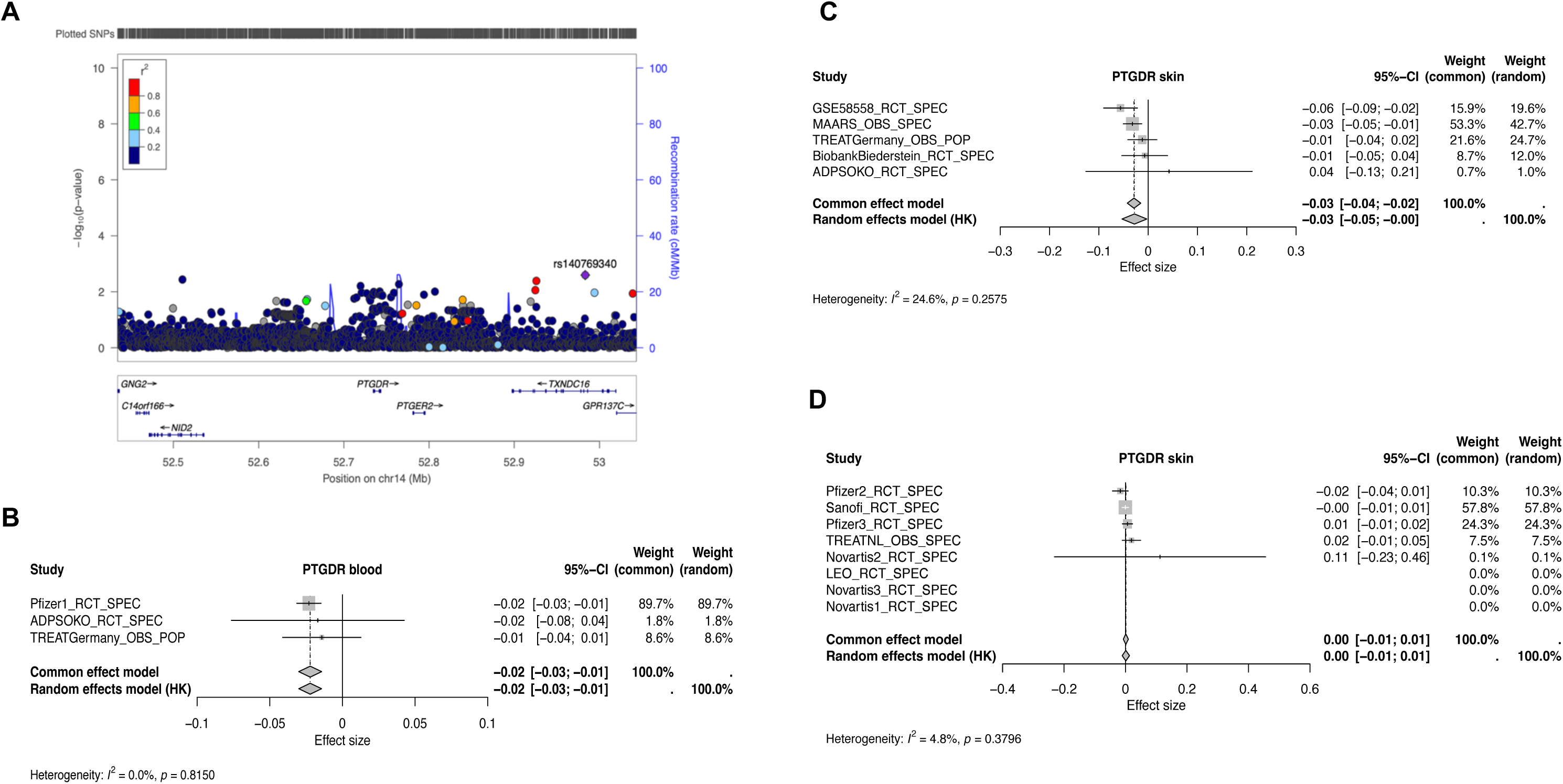
Evidence for *PTGDR* association with AD severity. (A) GWAS locus zoom; (B) association with blood transcriptomics; (C) association with skin transcriptomics; (D) independent replication of skin transcriptomic association.

**Supp Figure 7.**
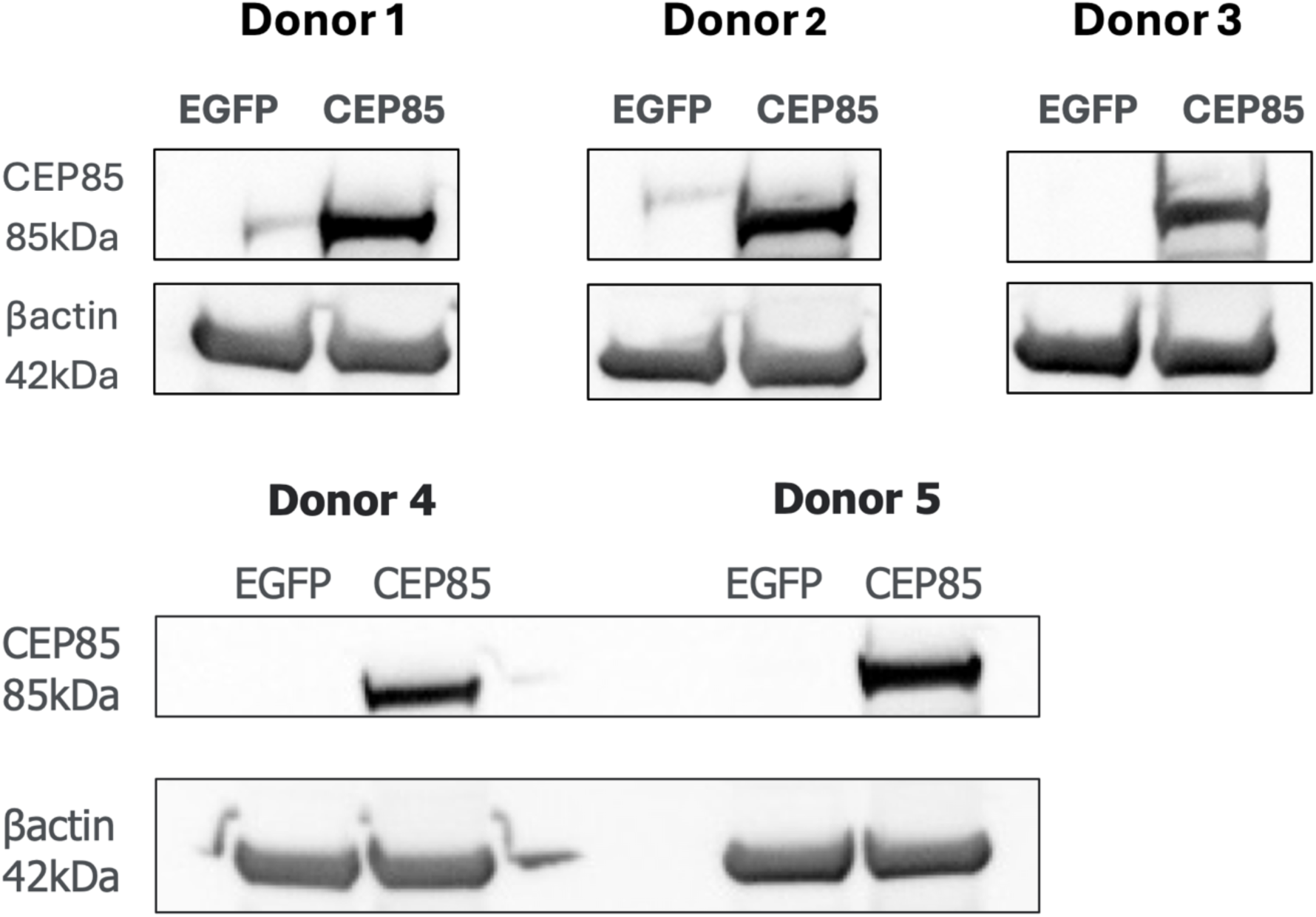
CEP85 western blot. Western blot approximately 18h post-transfection demonstrates successful overexpression of CEP85 in macrophages following mRNA transfection.

